# Estimation of time-varying causal effects with multivariable Mendelian randomization: some cautionary notes

**DOI:** 10.1101/2022.03.16.22272492

**Authors:** Haodong Tian, Stephen Burgess

## Abstract

**Introduction:** For many exposures present across the lifecourse, the effect of the exposure may vary over time. Multivariable Mendelian randomization (MVMR) is an approach that can assess the effects of related risk factors using genetic variants as instrumental variables. Recently, multivariable Mendelian randomization has been used to estimate the effects of an exposure during distinct time periods.

**Methods:** We investigated the behaviour of estimates from MVMR in a simulation study for different time-varying causal scenarios. We also performed an applied analysis to consider how MVMR estimates of body mass index on systolic blood pressure vary depending on the time periods considered.

**Results:** Estimates from MVMR in the simulation study were close to the true values when the outcome model was correctly specified: that is, when the outcome was a discrete function of the exposure at the precise timepoints at which the exposure was measured. However, in more realistic cases, MVMR estimates were not only incorrect, but misleading. For example, in one scenario MVMR estimates for early life were clearly negative despite the true causal effect being constant and positive. In the applied example, estimates were highly variable depending on the time period in which genetic associations with the exposure were estimated.

**Conclusions:** The poor performance of MVMR to study time-varying causal effects can be attributed to model misspecification and violation of the exclusion restriction assumption. We would urge caution about quantitative conclusions from such analyses and even qualitative interpretations about the direction, or presence or absence of a causal effect during a given time period.

## Introduction

Multivariable Mendelian randomization is an extension of standard (univariable) Mendelian randomization to investigate the causal effects of related risk factors with shared genetic predictors [1]. In standard Mendelian randomization, we take genetic variants that are predictors of a single risk factor, and assess whether genetically-predicted levels of the risk factor are associated with the outcome [2]. If the genetic variants satisfy the instrumental variable assumptions, then an association between genetically-predicted levels of the risk factor and the outcome is indicative of a causal effect of the risk factor on the outcome [3]. Multivariable Mendelian randomization is analogous, except that instead of testing whether genetically-predicted values of a single risk factor are associated with the outcome, we test whether genetically-predicted values of multiple risk factors are associated with the outcome or not in a multivariable model [4]. Estimates from multivariable Mendelian randomization represent direct effects of the risk factors on the outcome [5]. The method is primarily used in two contexts: first, to assess the effect of an exposure when genetic variants associated with the exposure may have pleiotropic effects on the outcome via other measured risk factors [1]; and second, to assess the relative contribution of causal pathways from the exposure to the outcome via other measured risk factors in a mediation analysis [5, 6].

We investigate a related application of multivariable Mendelian randomization that has been considered in empirical investigations [7, 8], and has recently been described from a methodological perspective [9, 10] that we refer to as time-varying Mendelian randomization. In this setting, the risk factors are not separate exposures, but rather multiple measures of the same exposure at different timepoints (Figure 1). For instance, Richardson *et al* considered body mass index (BMI) measured during early life and during later life as separate risk factors, and assessed whether genetically-predicted values of early life and later life BMI were associated with coronary artery disease (CAD) risk [7]. The authors interpreted a positive univariable association between genetically-predicted early life BMI and CAD risk as evidence that early life BMI is a causal risk factor for CAD, and lack of an independent association between genetically-predicted early life BMI and CAD risk in a multivariable model that also included genetically-predicted later life BMI as evidence that early life BMI does not have a direct effect on CAD risk, but that the risk is mediated via later life BMI.

**Figure 1:**
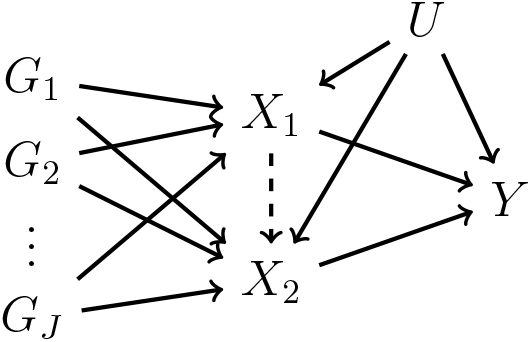
Directed acyclic graph illustrating multivariable Mendelian randomization assumptions for two risk factors (*X*_1_ and *X*_2_), *J* genetic variants (*G*_1_, *G*_2_, …, *G*_*J*_) that are assumed to satisfy the multivariable instrumental variable assumptions, an outcome (*Y*), and an unmeasured confounder (*U*). In time-varying Mendelian randomization, we assume that the two risk factors are measurements of the same exposure at different timepoints. The dashed arrow from *X*_1_ to *X*_2_ indicates the potential causal effect of the exposure at timepoint 1 on the exposure at timepoint 2.

These analyses pose several difficulties. First, it is necessary to have some linear independence between genetic predictors of the exposure at different timepoints [11]. If genetic associations with the exposure at different timepoints are perfectly proportional, then it is not possible to disentangle the effects of the exposure at different timepoints. It is not necessary to find distinct genetic predictors of the exposure at different timepoints, but if correlations across variants between the genetic associations with the exposure at different timepoints are strong, then the analysis will have limited utility.

However, even when such genetic variants are available, the use of multivariable Mendelian randomization to investigate time-varying causal effects necessitates strong parametric assumptions that are unlikely to be plausible in practice. In univariable Mendelian randomization, in order to estimate a parameter that represents a causal effect, it is necessary to make parametric assumptions [12]. However, even if these assumptions are not satisfied, the standard Mendelian randomization estimate is a valid test statistic for the sharp causal null hypothesis that the exposure has no causal effect on the outcome at any timepoint [13]. Rejection of this null hypothesis implies that the exposure has a causal effect on the outcome at some timepoint in at least a subset of the population. Here, we will show by counterexample that an analogous result for time-varying Mendelian randomization does not hold.

In this paper, we will explore the behaviour of estimates from multivariable Mendelian randomization for a time-varying exposure in a simple simulation study. We shall show that, when the model for the outcome is correctly specified, estimates from multivariable Mendelian randomization are close to the true parameter values. However, when the model is incorrectly specified (as is likely in practice), estimates are not only incorrect, but they can be highly misleading. We also perform an applied multivariable Mendelian randomization analysis of body mass index (BMI) on systolic blood pressure (SBP), and show that estimates and inferences are highly variable depending on the time period at which genetic associations with BMI are estimated.

## Methods

Multivariable Mendelian randomization has been described at length in the literature [1, 4], and in particular in the context of time-varying causal effects [9, 10]. We here provide a brief overview of the approach.

### Assumptions and estimation

We use the term ‘exposure’ to describe the putative causal factor, and the term ‘risk factor’ to describe the value of the exposure at a particular timepoint. Multivariable Mendelian randomization requires each genetic variant to satisfy the multivariable instrumental variable assumptions [14, ch. 9]:

i. the variant is associated with one or more of the risk factors,
ii. the variant is not associated with the outcome via a confounding pathway, and
iii. the variant does not affect the outcome directly, only possibly indirectly via one or more of the risk factors.

In order to estimate causal effects, we need to make additional assumptions. For simplicity, we assume that all variables are continuous, and the associations between the genetic variants and the risk factors, the genetic variants and the outcome, and the causal effect of the risk factors on the outcome are homogeneous and linear without effect modification by any confounder [3]. This enables the estimation of the direct effects of the risk factors on the outcome using the two-stage least squares (2SLS) method, which can be implemented by first regressing the risk factors on the genetic variants, and then regressing the outcome on the fitted values of the risk factors in a multivariable model [15, ch. 15].

The term ‘direct effect’ is imprecise; it has previously been argued that the direct effects estimated in instrumental variable analyses are most naturally interpreted as controlled direct effects (that is, the effect of varying the exposure while fixing the mediator at a given value), as the instrumental variables set the values of the risk factors [6]. However, in the all-linear setting, natural and controlled direct effects take the same value, so any difference is a question of interpretation. An alternative estimation method that could be applied here is g-estimation of structural mean models; although estimates are similar to those from 2SLS in the all-linear setting [10]. Equivalent estimates to those from 2SLS would also be obtained from the multivariable inverse-variance weighted method if we had access to summarized data on the genetic associations with the outcome and with the exposure at the relevant timepoints [16].

### Simulation studies

We investigate the behaviour of multivariable Mendelian randomization estimates for a time-varying exposure in a series of simulation studies. We simulate the time-varying exposure *X*(*t*) according to the following data-generating model:

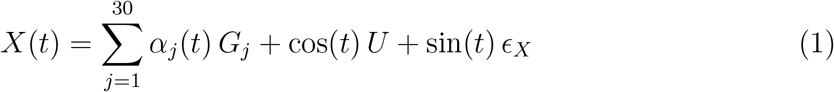

where the genetic variants *G*_*j*_ for *j* = 1, 2, …, 30, follow independent binomal distributions *B*(2, 0.3), and the confounder *U* and the error term *ϵ*_*X*_ have independent standard normal distributions. The exposure varies over time *t*, which can be interpreted as an individual’s age, owing to sinusoidal effects of the error term (sin(*t*)), the confounder (cos(*t*)), and the genetic variants:

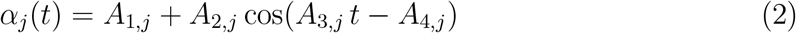

where *A*_1,*j*_, *A*_2,*j*_, *A*_3,*j*_, and *A*_4,*j*_ all have independent normal distributions for each variant *j*. The distributions of {*A*_1,*j*_, *A*_2,*j*_, *A*_3,*j*_, *A*_4,*j*_} are fixed for each scenario, but vary between scenarios. Detailed values for the independent normal distribution are shown in Supplementary Table S1.

We consider two scenarios for the outcome. First, we assume that the outcome is a function of the exposure at two fixed time-points, *t* = 10 and *t* = 50:

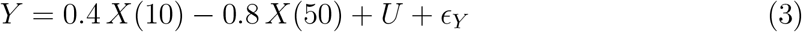

where *U* is the confounder as above and *ϵ*_*Y*_ has an independent standard normal distribution. The true causal effect of the exposure at time 10 is +0.4, and the causal effect of the exposure at time 50 is −0.8. We consider estimates from multivariable Mendelian randomization in four cases: first, when the exposure is measured at times 10 and 50 (Scenario 1A); then when the exposure is measured at times 10, 40, and 50 (Scenario 1B); at times 15 and 30 (Scenario 1C); and at times 15 and 50 (Scenario 1D). We recognize that this outcome model is somewhat unrealistic; we explore these scenarios to investigate whether methods can consistently estimate causal effects when the model is correctly specified.

Secondly and more realistically, we assume that the outcome is a continuous function of the exposure that varies over time. We express the relationship between the exposure and outcome using an integral, where the causal effect of the exposure depends on the time-varying function *β*(*t*):

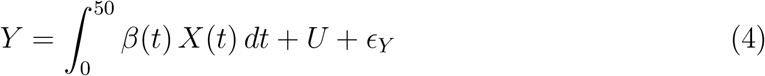

where again the confounder *U* and *ϵ*_*Y*_ have independent standard normal distributions. We consider three different scenarios for *β*(*t*):

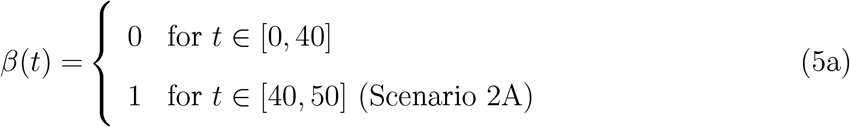

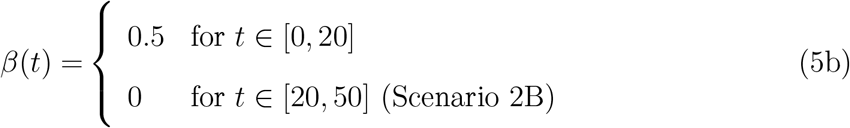

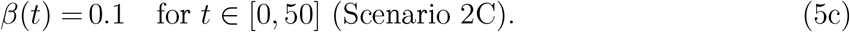

In Scenario 2A, the causal effect of the exposure is null in early life (up to time 40), and positive in later life. In Scenario 2B, the causal effect of the exposure is positive in early life (up to time 20), and null in later life. In Scenario 2C, the causal effect of the exposure is constant and positive across the life course. We consider multivariable Mendelian randomization estimates when the risk factors are the exposure measured at times 10 and 50. In Scenario 2C, we also consider a wider range of choices of timings for the exposure measurements, and consider the impact on estimates.

### Illustrative example: body mass index and systolic blood pressure

To investigate how estimates may behave in a real data analysis, we performed a time-varying multivariable Mendelian randomization analysis where the exposure is BMI and the outcome is SBP. Previous Mendelian randomization analyses have suggested that BMI has a positive causal effect on SBP [17], although how this effect may vary over time has not been explored. We took data from UK Biobank, a prospective cohort study of around half a million people aged 40 to 69 years at baseline, recruited in 2006-2010 from across the United Kingdom [18]. We considered 366,089 unrelated individuals of European ancestries, who passed various quality control filters as described previously [19]. We used 93 uncorrelated (pairwise *r*^2^ *<* 0.01) single nucleotide polymorphisms (SNPs) as instrumental variables which have previously been shown to be associated with BMI at a genome-wide level of statistical significance [20]. This genome-wide association study did not include UK Biobank participants, thus avoiding bias due to sample overlap [21].

We derived multivariable Mendelian randomization estimates using a two-sample 2SLS method, as we only have measurements of BMI for each individual at a single time point. In the first stage, we performed two separate regressions of BMI on the genetic variants using individuals recruited between ages 41-46 and 60-65. We used coefficients from these regressions to estimate genetically-predicted BMI at ages 41-46 and 60-65 for individuals aged *>* 65 at recruitment. We then performed the second-stage regression of SBP on genetically-predicted BMI using individuals aged *>* 65 at recruitment. To investigate variability of results to the choice of time period, we repeated this analysis using different age ranges for the first BMI regression: 42-47, 43-48, 44-49, and so on up to 55-60. The second time period was always taken as 60-65, and the second-stage regression was always conducted in individuals aged *>* 65 at recruitment. Estimates represent the change in SBP in mmHg per 1 kg/m2 increase in genetically-predicted BMI.

## Results

### Simulation study

For each scenario, we generated 1000 datasets on 10 000 participants. The average proportion of variance in the exposure explained by the genetic variants under each scenario is around 10%, corresponding to a univariable F statistic of 36.9. The average conditional F statistic values for multivariable Mendelian randomization in most scenarios are larger than 10, a value conventionally regarded as a guide to diagnose weak instruments [4, 11]. Detailed values for instrument strength values are shown in Supplementary Table S2. We present results from multivariable Mendelian randomization obtained using the 2SLS method.

### Discrete effects of the exposure at specific time-points

Results from the simulation study where the outcome is a function of the exposure at times 10 and 50 are shown in Figure 2. In Scenario 1A, we use the exposure measured at times 10 and 50 as risk factors. Median estimates across scenarios are close to the true causal effects: namely +0.4 at time 10 and −0.8 at time 50, though there is some bias towards the null due to weak instruments. In Scenario 1B, we use the exposure measured at times 10, 40, and 50 as risk factors. Again, median estimates across scenarios are close to the true causal effects, with the median estimate at time 40 around zero. However, in Scenario 1C (using the exposure at times 15 and 30) and in Scenario 1D (using the exposure at times 15 and 50), median estimates are substantially different to the true values. In Scenario 1C, the median estimate at time 15 is negative and at time 30 is positive; this is the opposite to the true situation, as the true effect is positive at the earlier time point and negative at the later timepoint. In Scenario 1D, the median estimate at time 50 is correctly negative, but the median estimate at time 15 is also negative, while the early-age causal effect is positive. We repeated the simulation study with stronger instruments (average proportion of variance in the exposure explained was around 30%); results shown in Supplementary Figure S1 support our claim that bias in Scenarios 1A and 1B is due to weak instruments.

**Figure 2:**
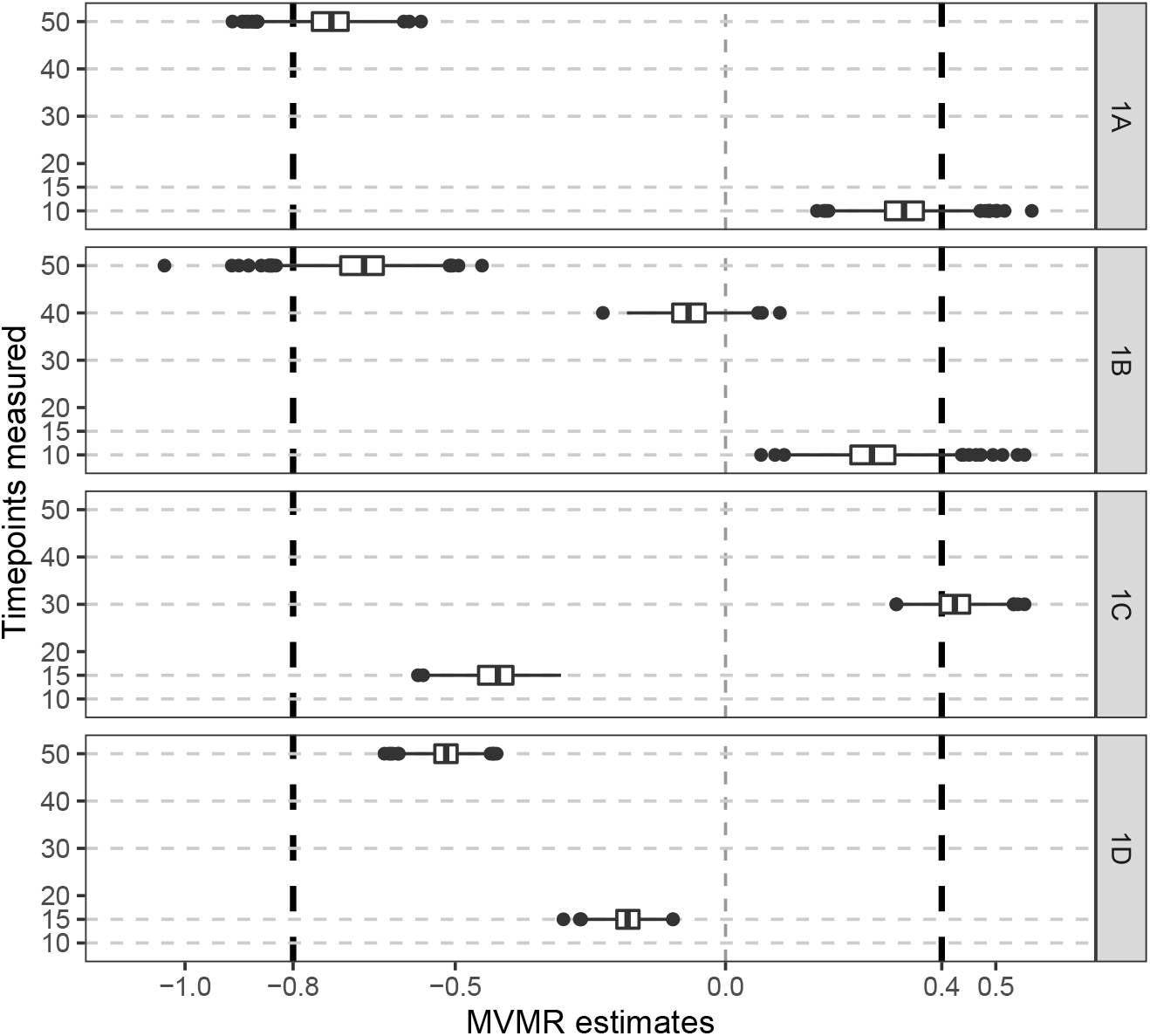
Simulation results when the outcome is affected discretely by the exposure at specific time-points. Boxplots of multivariable Mendelian randomization (MVMR) estimates with risk factors taken as the exposure measure at different measured timepoints – Scenario 1A: times 10 and 50; Scenario 1B: 10, 40, 50; Scenario 1C: 15, 30; Scenario 1D: 15, 50. Box indicates lower quartile, median, and upper quartile; error bars represent the minimal and maximal data point falling in the 1.5 interquartile range distance from the lower/upper quartile; estimates outside this range are plotted separately. The true effects are *β*_1_ = 0.4 at time 10 and *β*_2_ = −0.8 at time 50 (black dashed lines).

### Continuous effects of the exposure across time

Results from the simulation study where the outcome is a continuous function of the exposure are shown in Figure 3. In Scenario 2A, median estimates at both timepoints are positive despite the true effect being null in early life and positive in later life only. In Scenario 2B, the median estimate at the earlier timepoint is close to zero and the median estimate at the later timepoint is positive, whereas in truth the causal effect of the exposure is positive in early life and null in later life. In Scenario 2C, the median estimate at the earlier timepoint is negative and the median estimate at the later time-point is positive, despite the true causal effect of the exposure being constant and positive throughout.

**Figure 3:**
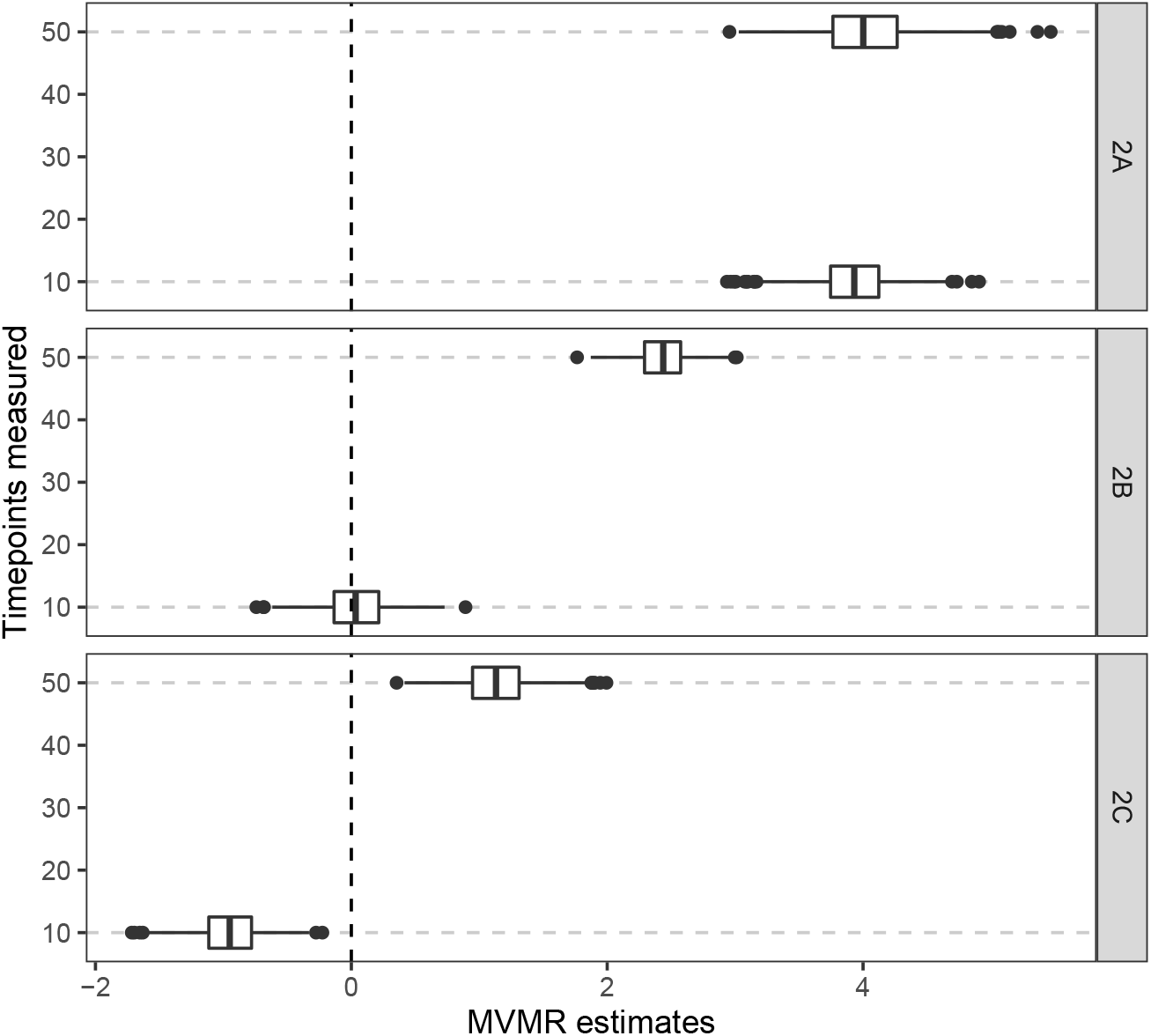
Simulation results when the outcome is affected continuously by the exposure across time. Boxplots of multivariable Mendelian randomization (MVMR) estimates with risk factors taken as the exposure measure at different measured timepoints: Scenario 2A: null effect until time 40, positive effect thereafter; Scenario 2B: positive effect until time 20, null effect thereafter; Scenario 2C: constant positive effect. Box indicates lower quartile, median, and upper quartile; error bars represent the minimal and maximal data point falling in the 1.5 interquartile range distance from the lower/upper quartile; estimates outside this range are plotted separately.

Figure 4 provides results from Scenario 2C (constant positive effect) for a range of different choices of timepoints, indicated as Option 1, Option 2, and so on. We also vary the number of timepoints considered. Median estimates varied substantially depending on the choice of timepoints, with no evident pattern of results. Although median estimates were mostly positive, they were sometimes close to zero and occasionally clearly negative. Moreover, it is not clear that estimates improved when considering measures of the exposure at increased numbers of timepoints, as some median estimates were either close to zero or negative even when the exposure was measured at four timepoints.

**Figure 4:**
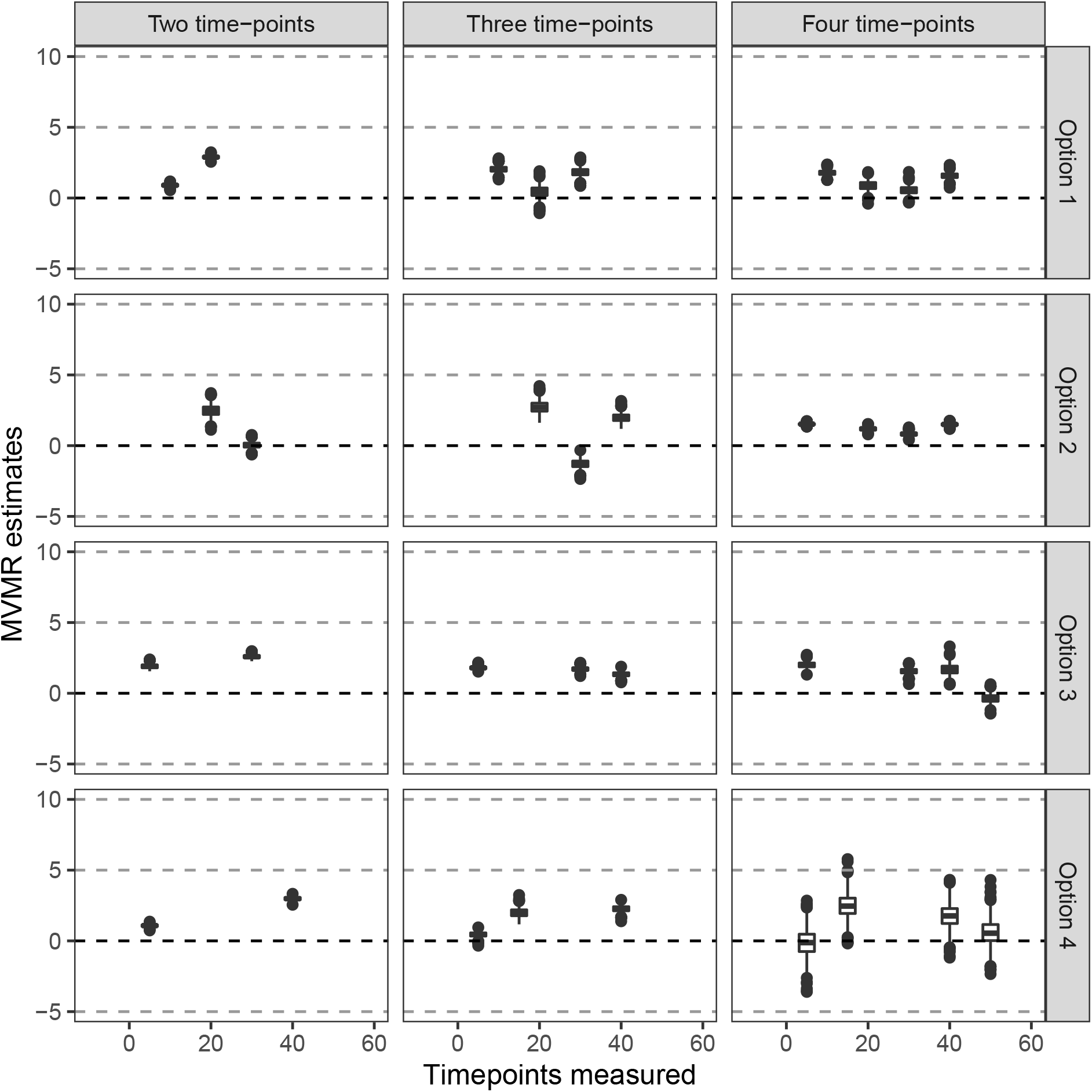
Simulation results when the outcome is affected continuously by the exposure across time in Scenario 2C (constant positive effect). Boxplots of multivariable Mendelian randomization (MVMR) estimates with risk factors taken as the exposure measure at different measured timepoints when varying the location and number of timepoints. Box indicates lower quartile, median, and upper quartile; error bars represent the minimal and maximal data point falling in the 1.5 interquartile range distance from the lower/upper quartile; estimates outside this range are plotted separately.

### Illustrative example: body mass index and systolic blood pressure

Results from the illustrative example for the effect of BMI on SBP are shown in Figure 5. Scatter plots of the genetic associations with BMI at different time periods are shown in Supplementary Figure S2; while most variants are similarly associated with BMI across time periods, some variants are more strongly associated during the earlier or later time period, enabling the multivariable Mendelian randomization analysis to be performed. Multivariable Mendelian randomization estimates for both time periods are highly variable about the age range over which the risk factor associations were estimated. The range of variation in Mendelian randomization estimates was greater than expected based on the standard errors of estimates. In some cases, the 95% confidence interval for the estimate in the first time period was negative and excluded the null and the 95% confidence interval for the estimate in the second time period was positive and excluded the null. But there were also positive point estimates for the first time period and negative point estimates for the second time period. In summary, both estimates and inferences based on those estimates were strongly dependent on the choice of the first time period, a choice that is likely to be arbitrary in practice.

**Figure 5:**
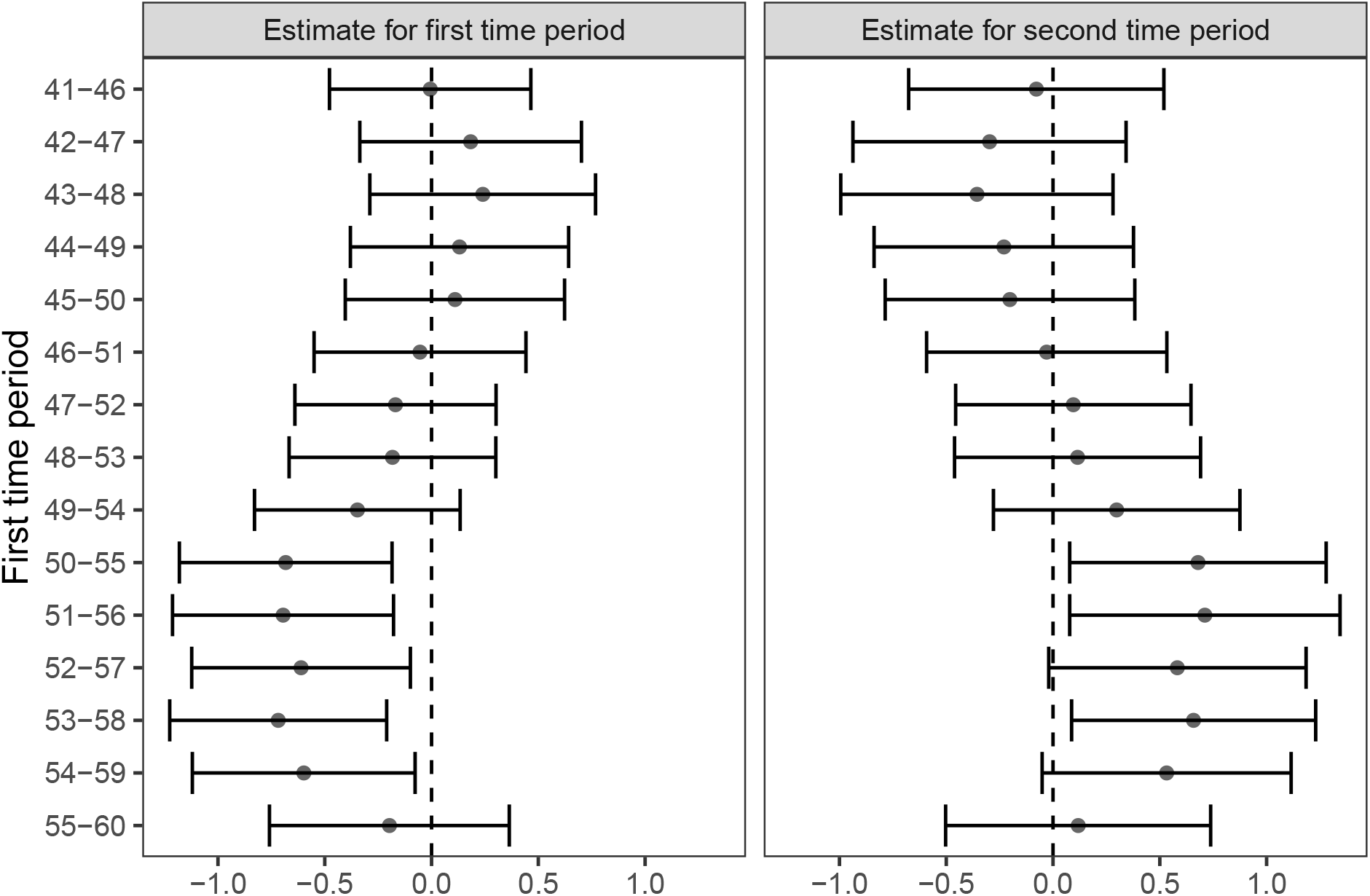
Multivariable Mendelian randomization estimates (95% confidence intervals) of the effect of body mass index (BMI) on systolic blood pressure for different time periods. The first risk factor is BMI over the first time period as indicated. The second risk factor is BMI over the time period from age 60 to 65. Estimates for systolic blood pressure are performed in individuals aged *>* 65 at the time of recruitment.

## Discussion

As the well-known aphorism goes: all models are incorrect, but some models are useful [22]. In univariable Mendelian randomization, strict parametric assumptions are required for Mendelian randomization estimates to be interpreted as either an average causal effect or a local average causal effect [12]. However, even if these assumptions are not satisfied, the univariable Mendelian randomization estimate still has an interpretation as a test statistic for a relevant causal hypothesis [13]. Indeed, it is been argued that the numerical value of a Mendelian randomization estimate rarely represents the estimate of a policy-relevant parameter [23], as (amongst other reasons) it represents the impact of a lifelong change in the distribution of the exposure, whereas interventions on exposures in clinical practice are more limited in time [24, 25]. Hence the primary value of a Mendelian randomization investigation is to provide evidence supporting (or questioning) a causal hypothesis, rather than providing a causal estimate [26].

However, estimates from multivariable Mendelian randomization for a time-varying exposure do not seem to have a similar interpretation as a test statistic for a relevant causal hypothesis relating to the presence of a causal effect over a specific time period. When the exposure affects the outcome at a limited number of discrete timepoints and the risk factors in the multivariable Mendelian randomization analysis are the values of the exposure at these timepoints, causal effects at these timepoints can be unbiasedly estimated. But if these timepoints are not correctly identified, estimates are not only biased, they are incorrect in a way that is misleading to any inferences being drawn from their magnitude, either about the presence or the direction of a causal effect. Similarly, if the effect of the exposure on the outcome is not discrete, but rather continuous, then estimates can be biased in a misleading way. It is implausible for the true effect of an exposure on the outcome to be discrete at the precise timepoints at which measurements of the exposure are available; hence, we expect potentially misleading bias to be ubiquitous for time-varying Mendelian randomization investigations in practice.

We have provided a simulated example where the effect of the exposure on the outcome is positive in early life only and negative in later life, but estimates from multivariable Mendelian randomization would suggest the opposite. We have also provided a simulated example where the effect of the exposure on the outcome occurs in early life only and not in later life, but estimates from multivariable Mendelian randomization would suggest the opposite. Similarly, we have provided a simulated example where the effect of the exposure on the outcome occurs in later life only and not in early life, but estimates from multivariable Mendelian randomization analysis are positive throughout. Finally, we have provided a simulated example where estimates from multivariable Mendelian randomization at different timepoints are not necessarily positive even though the effect of the exposure is uniformly constant and positive. We have also provided an applied example with real data, and shown that estimates are sensitive about the time period over which genetic associations with the exposure were estimated. The choice of time period influenced the conclusions drawn from the analysis: whether there is a negative effect of BMI on SBP during the first time period or not, and whether there is a positive effect of BMI on SBP during the second time period or not.

The poor performance of multivariable Mendelian randomization in these examples can be attributed to model misspecification. In general, when a statistical model is misspecified, estimates suffer from bias that is unpredictable in both magnitude and direction [27]. Another explanation is violation of the exclusion restriction assumption. The instrumental variable assumptions state that the totality of the effect of the genetic variants on the outcome is mediated via the exposure, such that if the exposure were fixed and the genetic variants varied, the outcome would remain the same [28]. However, if the effect of the exposure acts continuously over time, then it would not be sufficient to fix its value at a few specific timepoints, but it would be necessary to fix the trajectory of the exposure over time. Similarly, complete mediation of the genetic effect on the outcome via the exposure would only be observed when considering the distribution of the exposure over time. As time is continuous rather than discrete, it is unlikely that any model containing values of the exposure at specific discrete timepoints could be correctly specified. One potential extension of this work is to consider models that fit the outcome as a continuous function of the exposure.

One qualitative limitation of our cautionary remarks is that we have considered scenarios in which the risk factors represent the same exposure measured at different time-points. In the example of Richardson *et al* [7], it is arguable that early life BMI and later life BMI represent distinct exposures, and hence multivariable Mendelian randomization is a legitimate tool for obtaining meaningful inferences in this case. While we agree that such a situation is more amenable to the use of multivariable Mendelian randomization, we would still be cautious about overinterpretation of results from time-varying Mendelian randomization investigations, and would recommend that investigators assess the robustness of findings carefully (for example, by assessing the consistency of results with different choices of genetic variants and/or different timepoints for the measurement of the exposure). Another limitation is that the simulation scenarios considered may be unrealistic, although similar fragility in estimates was observed in the applied analysis.

In conclusion, multivariable Mendelian randomization analyses to investigate time-varying causal effects rely on parametric assumptions that are unlikely to be satisfied in practice, and provide estimates that can be misleading if the model is incorrect. We therefore strongly discourage quantitative conclusions to be drawn from these analyses, and would advise caution about the qualitative interpretation of such findings as a guide of the direction or even existence of a causal effect during a particular phase of life.

## Data Availability

All data produced in the present study are available upon reasonable request to the authors

## Ethical approval

The UK Biobank study has approval from the North West Multicentre Research Ethics Committee (11/NW/0382). The research has been conducted using the UK Biobank Resource under Application Number 7439.

## Author contributions

HT designed and conducted the simulation and applied study. SB directed the study’s implementation and helped to interpret the findings. All authors reviewed and edited the manuscript.

## Acknowledgements

We thank Christopher Jackson and Jessica Barrett (MRC Biostatistics Unit) for helpful discussions in the development of this work. This research was conducted using the UK Biobank Resource under Application Number 7439.

## Supplementary Material

**Supplementary Table S1:** The independent normal distribution used in the effect parameters (*A*_1_, *A*_2_, *A*_3_, *A*_4_ in (2)) of genetic variants for each simulation study scenario. N(*µ, σ*^2^) representing the normal distribution with the mean *µ* and the variance *σ*^2^.

| Scenario | $A_1$ | $A_2$ | $A_3$ | $A_4$ |
| --- | --- | --- | --- | --- |
| 1 | $\mathcal{N}(0, 0.05^2)$ | $\mathcal{N}(0, 0.15^2)$ | $\mathcal{N}(3, 0.01^2)$ | $\mathcal{N}(0, 1^2)$ |
| 1 (with stronger instruments) | $\mathcal{N}(0, 0.1^2)$ | $\mathcal{N}(0, 0.3^2)$ | $\mathcal{N}(3, 0.01^2)$ | $\mathcal{N}(0, 1^2)$ |
| 2A | $\mathcal{N}(0, 0.05^2)$ | $\mathcal{N}(0, 0.05^2)$ | $\mathcal{N}(0, 1^2)$ | $\mathcal{N}(0, 1^2)$ |
| 2B | $\mathcal{N}(0, 0.05^2)$ | $\mathcal{N}(0, 0.15^2)$ | $\mathcal{N}(3, 0.01^2)$ | $\mathcal{N}(0, 1^2)$ |
| 2C | $\mathcal{N}(0, 0.05^2)$ | $\mathcal{N}(0, 0.15^2)$ | $\mathcal{N}(0.1, 0.01^2)$ | $\mathcal{N}(0, 1^2)$ |

**Supplementary Table S2:** Average values of instrument strength in the various simulation study scenarios. *R*^2^ represents the average proportion of variance in the exposure explained by the genetic variants. F and conditional F is the corresponding univariate and conditional F statistic value.

| Scenario | timepoint | $R^2$ | F | conditional F |
| --- | --- | --- | --- | --- |
| 1A | 10 | 0.099 | 36.4 | 19.9 |
|  | 50 | 0.106 | 39.2 | 21.5 |
| 1B | 10 | 0.099 | 36.4 | 13.0 |
|  | 40 | 0.129 | 49.4 | 29.7 |
|  | 50 | 0.106 | 39.2 | 13.6 |
| 1C | 15 | 0.120 | 45.2 | 33.5 |
|  | 30 | 0.147 | 57.3 | 42.4 |
| 1D | 15 | 0.120 | 45.2 | 45.1 |
|  | 50 | 0.106 | 39.2 | 35.2 |
| 2A | 10 | 0.038 | 13.2 | 11.4 |
|  | 50 | 0.027 | 9.1 | 7.9 |
| 2B | 10 | 0.116 | 43.5 | 27.9 |
|  | 50 | 0.157 | 62.0 | 39.8 |
| 2C | 10 | 0.097 | 35.8 | 20.2 |
|  | 50 | 0.086 | 31.5 | 17.8 |

**Supplementary Figure S1:**
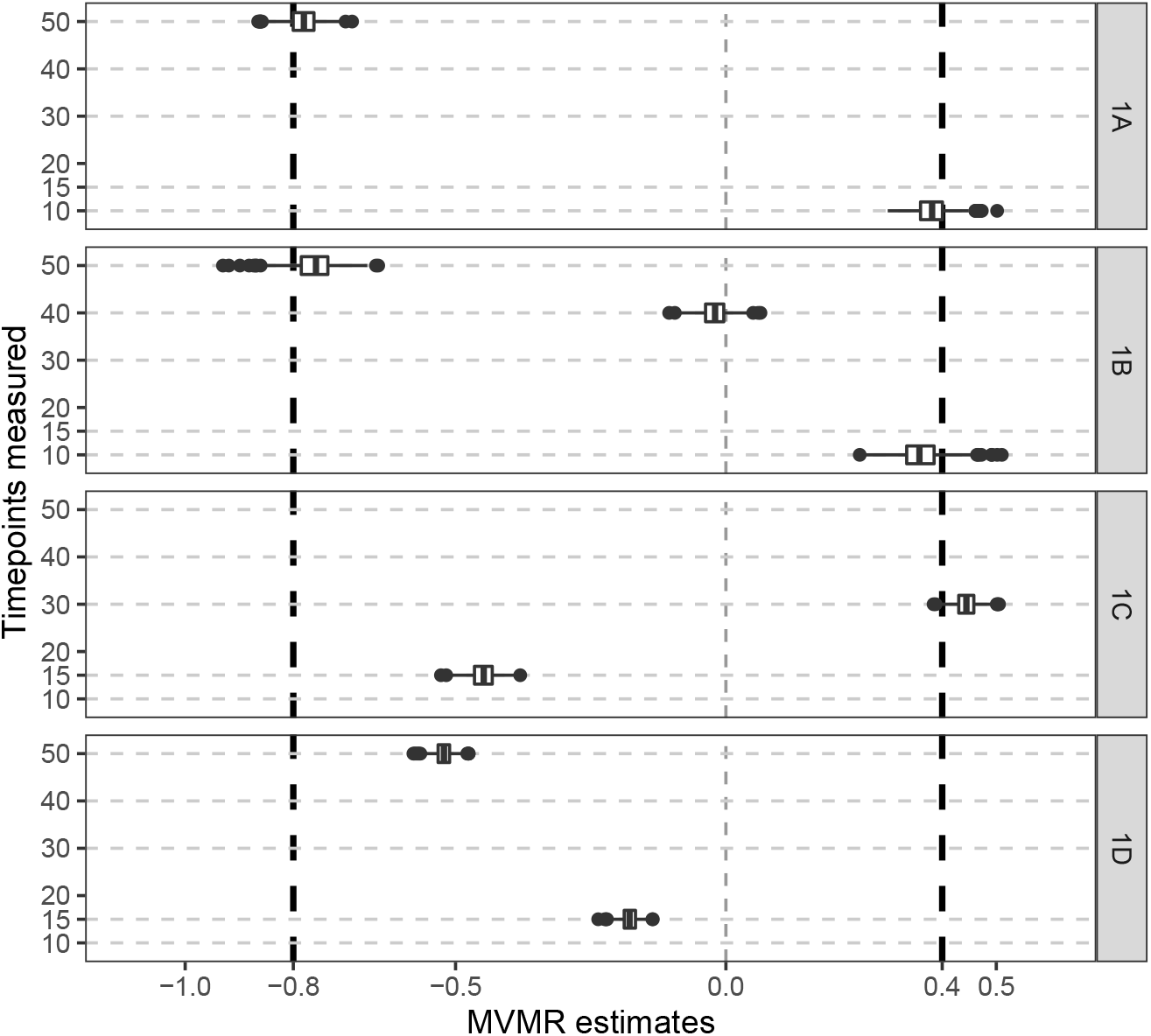
Simulation results with stronger instruments (the average proportion of variance in the exposure explained was around 30%) when the outcome is affected discretely by the exposure at specific time-points. Boxplots of multivariable Mendelian randomization (MVMR) estimates with risk factors taken as the exposure measure at different measured timepoints – Scenario 1A: times 10 and 50; Scenario 1B: 10, 40, 50; Scenario 1C: 15, 30; Scenario 1D: 15, 50. Box indicates lower quartile, median, and upper quartile; error bars represent the minimal and maximal data point falling in the 1.5 interquartile range distance from the lower/upper quartile; estimates outside this range are plotted separately. The true effects are *β*_1_ = 0.4 at time 10 and *β*_2_ = − 0.8 at time 50 (black dashed lines).

**Supplementary Figure S2:**
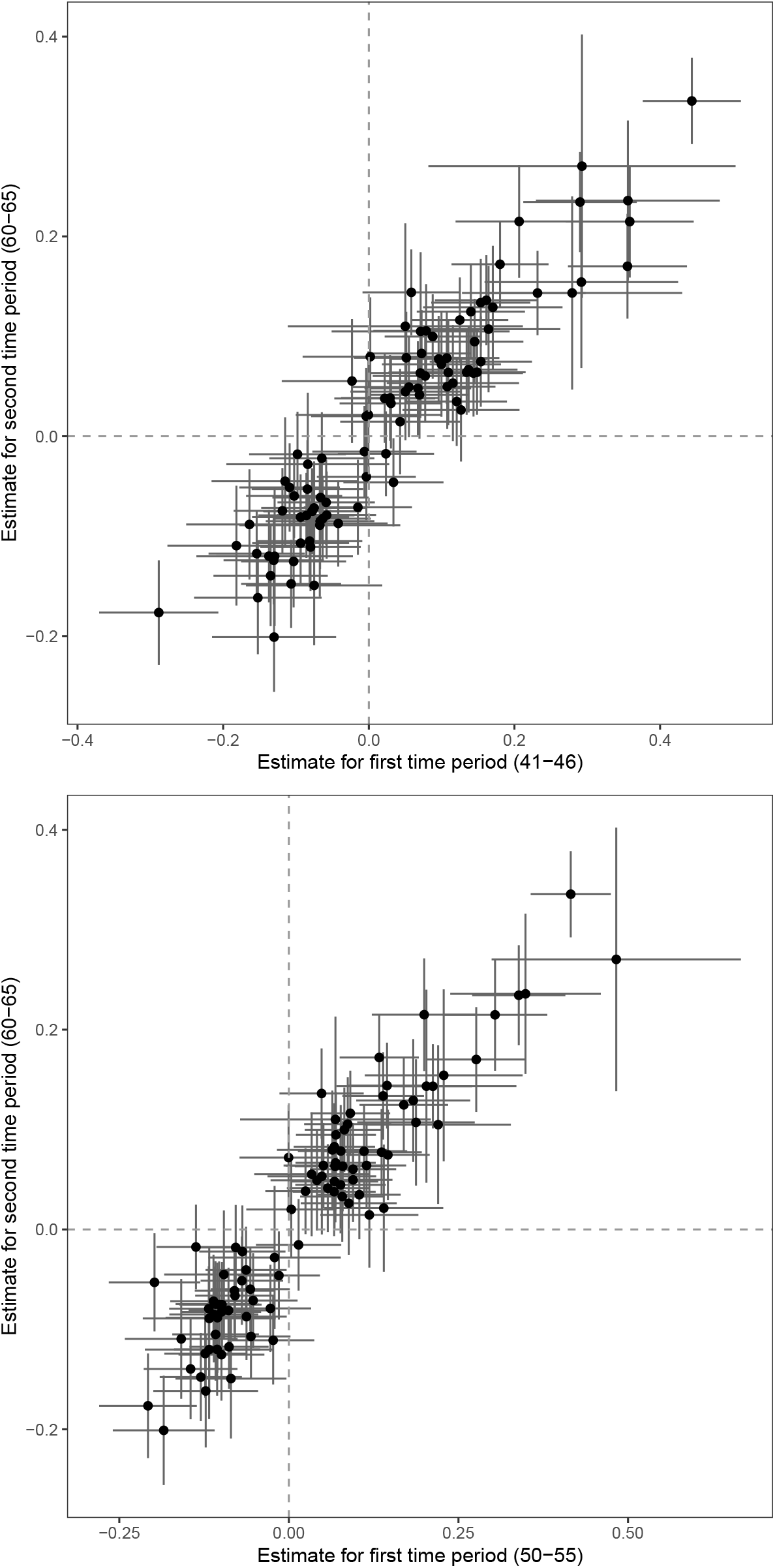
Scatter plots of the instrument-exposure associations at two different time periods for 93 genetic variants. In the top case, the two time periods in which the exposure was measured are 41-46 and 60-65. In the bottom case, the two time periods are 50-55 and 60-65. The error bar represents the 95% confidence interval for the corresponding instrument-exposure association estimate.

